# Debiasing Covid-19 prevalence estimates

**DOI:** 10.1101/2021.01.10.21249298

**Authors:** Sotiris Georganas, Alina Velias, Sotiris Vandoros

## Abstract

Timely, accurate epidemic figures are necessary for informed policy. In the Covid-19 pandemic, mismeasurement can lead to tremendous waste, in health or economic output. “Random” testing is commonly used to estimate virus prevalence, reporting daily positivity rates. However, since testing is necessarily voluntary, all “random” tests done in the field suffer from *selection bias*. This bias, unlike standard polling biases, goes beyond demographical representativeness and cannot be corrected by oversampling (i.e. selecting people without symptoms to test). Using controlled, incentivized experiments on a sample of all ages, we show that people who feel symptoms are up to 33 times more likely to seek testing. The bias in testing propensities leads to sizable prevalence bias: test positivity is inflated by up to five times, even if testing is costless. This effect varies greatly across time and age groups, making comparisons over time and across countries misleading. We validate our results using the REACT study in the UK and find that positivity figures have indeed a very large and time varying bias. We present calculations to debias positivity rates, but importantly, suggest a parsimonious way to sample the population bypassing the bias altogether. Our estimation is both real-time and consistently close to true values. These results are relevant for all epidemics, besides covid-19, when carriers have informative beliefs about their own status.

## 1. Background

Tackling the covid-19 pandemic is of paramount importance for obvious health and financial reasons. Over 2 million deaths have been confirmed globally (Johns Hopkins 2020), and excess mortality is reported,^1^ as well as indirect health effects (undiagnosed/undertreated diseases^2-3^ and suicides.^4-5^). The pandemic per se, but also the containment measures against it, have crippled economic activity, leading to increasing unemployment rates and shrinking income worldwide^6^ – in turn leading to further deterioration of health.^7-8^ The design of optimal interventions to fight the disease, in time and space, requires efficient and accurate prevalence measurement, preferably in real time.

How to measure prevalence for infectious diseases? In this paper we show that commonly used methods are flawed because testing is voluntary, leading to heavy *self-selection bias*. We calculate how self-selection translates into *biased prevalence estimates* generally, and estimate the likely size of this bias, employing incentivised controlled experiments on a large sample of all ages, using standard experimental methods (similar were used in an influential paper to estimate HIV testing demand^9^, but not disease prevalence). Using our experimental data, we find that the commonly used “test positivity” measure may inflate actual prevalence by up to 5 times, even if testing is provided at zero cost. If covid-19 tests are costly for the testee (as is common), this inflation factor or *prevalence bias* can be much higher. To make estimation harder, the prevalence bias is *not constant*, but rather depends strongly on actual prevalence. This means that we cannot apply a fixed adjustment to test positivity measures, and such measures cannot be used to compare prevalence across countries, as is commonly done. To validate our results, we compare prevalence estimates from the REACT study in the UK, to test positivity ratios at the same dates^10^. As predicted by our calculations, the prevalence bias is indeed positive, very large and time varying, ranging from 3.8 to 23.6 in the different waves of the study. To say it another way, our estimates of the testing bias and calculations of how this translates to a prevalence bias, explain why test positivity rates always seem to be too high.

Our estimates of the testing bias by age group can be used to calculate the prevalence bias for countries with different demographic structures. Further, we present the parameter estimations necessary to debias the current prevalence estimates in the field, but, crucially, suggest a novel method to *bypass* the self-selection bias altogether with an estimation procedure that is at the same time faster, more accurate and more feasible than current methods.

To understand the relevance of these results, start by noting that suggested policy responses and their implementation (e.g. social distancing rules) will inevitably be inefficient if we are not aware of the real number of active cases, and in which areas and age groups these occur. Observing mortality rates or the number of hospitalisations and patients in ICU are not real time measurements; they only provide an estimate of how many people caught Covid-19 *weeks earlier* (and estimating the fatality rate is also challenging).^11^ This time lag is very important when trying to evaluate interventions. Without real time data, measuring the effect of a vaccine will take months, on top of the time the vaccine takes to have a medical effect. Understanding the full effect of other events on the disease, like the Christmas holidays (which led to more interaction and possibly higher transmission) similarly takes months (see excellent work on the effectiveness of NPIs, which uses death counts, lagging by several weeks)^12^. On the other hand, knowing the *current* number of actual cases, allows the design of optimal policy response, and also provides a forward-looking estimate of hospitalisations and mortality. Health systems get warning several weeks ahead, gaining invaluable time for necessary adjustments.

Community testing, often conducted in the high street and in neighbourhoods, is widely considered a useful tool to monitor incidence and trends. The ECDC^12^ lists “[to] reliably monitor SARS-CoV-2 transmission rates and severity” among five objectives of testing. It also publishes weekly testing data and “positivity rates” by EU State.^13^ However, we show that such testing cannot provide accurate estimates of Covid-19 prevalence, and the main problem is not related to typical issues that arise in population sampling, such as sampling representative age groups. Testing has to be voluntary, and people are likelier to self-select into testing if they have reasons to believe they might be having Covid-19 (such as, e.g. if they have symptoms or if they are exposed to a high-risk environment). We find in our data that the *self-selection bias* increases non-linearly with waiting times and any other cost associated with testing. To make prevalence estimation harder, the bias is time-varying, because it depends non-linearly on time varying parameters. For example, when cases rise steeply, people might be more likely to want to test out of fear. This leads to longer queues for testing, longer waiting times and a disproportionately larger testing bias.

To summarise, the objective of this paper is to examine whether and to what extent bias occurs in Covid-19 testing, to offer a debiasing solution to accurately estimate Covid-19 prevalence in the field using existing procedures, and lastly to propose a better procedure, both more economical and more accurate.

## 2. Data and Methods

Standard theoretical arguments allow the precise calculation of the prevalence bias (see Appendix C). However, the size and direction of the prevalence bias in the field is an empirical issue, and crucially depends on the self-selection testing bias based on symptoms. Are people who believe they have symptoms more likely to seek testing, and if yes, by how much? In order to measure this, we ran incentivised controlled experiments.

Data collection took place over a week, from 11 till 18 December 2020. The majority of the responses were collected online, via the *QualtricsTX* platform. To enable greater representativeness of the sample, 94 responses (16%) from elder people (median age = 63) were collected using phone interviews. Out of 608 participants starting the online study, 24 (4.7) dropped out mostly after the first few questions, resulting in the final sample of 578 observations.

The median age for the sample was 39 years (median for Greece 45.6), and the age distribution is shown in Figure A1 in Appendix A.

Subjects were invited to participate in a study on Covid-19 and related behaviours. Upon signing a consent form, the participant was first asked about general and Covid-19-related health. We then elicited hypothetical willingness to wait (WTW) to take a rapid test for Covid-19, *conditional* on (i) feeling healthy, (ii) having flu-like symptoms, (iii) having Covid-19 like symptoms. For all three hypothetical scenarios, the test was being offered by the national health authority (EODY) while the participant was walking down the street. This was done to reduce the (hypothetical) travel costs and reliability-related concerns.

After eliciting the hypothetical WTW, we asked the subjects several control questions, including exposure to Covid-19 risky environments (e.g. taking public transport or working fate-to-face with many people) and socio-demographics. After completing the compulsory part of the study, the participants were offered an optional task for which they were randomly allocated to one of the two prize treatments. In treatment *Book*, the participant would enter a 1/30 chance lottery for a voucher for the local large-scale bookshop chain (“Public”), worth €80. In treatment *Test*, the participant would enter the same 1/30 chance lottery for a voucher for a home-administered Covid-19 test. For both prizes, the delivery was guaranteed within the next 36 hours. All 578 participants completed the hypothetical elicitation and the control questions (left part of Figure A2 in Appendix A).

As was partly expected, a substantial part of the sample (n=174) did not continue to the optional task. A major part of it (n=78) was the elder people subsample. We are not very concerned that the inconvenience of the waiting task over the phone was the issue, since the participants came from the sample that previously participated in a study involving a real effort task over the phone^14^. For n=38 participants, a software glitch in Qualtrics, in the first five hours of the study resulted in missing recording of the treatment allocation, so we had to drop their data despite completion of the optional task.

The participants then read the description of the optional task. They learned that it involved waiting in front of their screen for some time (target) that would be revealed in the next screen, and the lottery draw for the prize would take place right after the wait. They also learned that to ensure that they are waiting, a button would appear at random times and they would need to press it within 4 seconds to avoid being disqualified. Among the 303 participants who read the description of the optional task, 241 continued to the next screen which revealed the waiting target. At this stage, they were randomly allocated to one of the four waiting target conditions {300, 600, 900, 1200}. Upon learning the wait time, further 59 participants dropped out instantly (median target time 900 seconds). Among the 241 waiting, 69 dropped out before completing the full wait (median target time 900 seconds). In total, 172 participants completed the waiting target (median target 600 seconds).

Upon completing the waiting task, each participant was randomly allocated to one of the four *Cash* conditions, {€20, €35, €50, €65}. The participant was offered a choice to enter the lottery for: (a) the original prize (*Book, Test*), or (b) the displayed *Cash* amount. Out of the 172 participants, 112 chose to swap the original prize for the cash amount, whilst 60 chose to stay with the original prize (median cash value €35 for both). A total of 7 participants won the lottery.

## 3. Results

Table 1 shows the ratio of willingness to test between people with symptoms and those without. The figure ranges between 1.5 and 38, depending on the age group and waiting times. People under 30 with symptoms are 1.532 times more likely to test when there is no waiting time, compared to those without symptoms. This figure increases to 2.882 when there is a short wait of 5-15 minutes; 4.423 with a 15-30 minute wait; 15.5 with a 30-60 minute wait and 38 with a 1-2 hour wait. The ratio for 30-50 year-olds rages between 1.517 for no wait and 16 for a 1-2 hour wait. For over 50-year-olds, the ratio ranges between 1.708 and 11.333. Overall, there is a bias even for no waiting time at all, which increases steeply for long waiting times in all age groups. Note that the bias also varies by other observable characteristics, for example, for waiting times of two hours and more, it is 84% higher for men than for women. Also, the propensity bias is 50% higher for obese people than for people in the healthy range, which indicates that people at risk not only have higher propensity to test (as is to be expected) but also react stronger to symptoms.

**Table 1.** Bias (‘survival’ ratio of people with covid-19 symptoms to people with no symptoms) by waiting time for rapid test, *N*=510

| Age group | No wait/<br>Immediate | 5-15 min | 15-30 min | 30-60min | 1-2h | over 2h | $N$ |
| --- | --- | --- | --- | --- | --- | --- | --- |
| Under 30 | 1.532 | 2.882 | 4.423 | 15.5 | 38 | 38 + | 181 |
| 30-50 | 1.517 | 3.102 | 5.8 | 8.857 | 16 | 16 + | 199 |
| 50+ | 1.708 | 2.706 | 3.571 | 11 | 11.333 | 11.333 + | 130 |
| Total | 1.564 | 2.918 | 4.567 | 11.2 | 17.143 | 17.143 + | 510 |

The *propensity to test bias* translates to a biased virus *prevalence estimate* (*β*) which is also time varying. Crucially it depends on symptom prevalence, which, given the exponential spread of Covid-19, can change massively in a short period of time. This means that the estimate depends on symptom prevalence, but the bias itself also depends on it – so the bias is time variant.

Apart from waiting times, self-selecting into testing also depends on the cost associated with it (if applicable – costs can vary from time to monetary value, travel etc). We found that the bias is associated with willingness to pay for the test (Table A2 in Appendix A). Of those who won a test voucher, 83.8% swapped it for cash, as opposed to 48.9% of those who won the book voucher, indicating that the majority of subjects would not be willing to pay to receive a test. However, the scope of this article is to correct bias for free tests subject to different waiting times, and further experiments are needed to reach concrete conclusions on willingness to pay.

We have launched an online calculator that provides estimates on the testing bias (available at http://georgana.net/sotiris/task/atten/covid.php). The bias calculations that lead to the formula on which the calculator is based, is provided in Appendix C. The estimates on the testing bias depend on (a) the percentage of tests yielding positive results; (b) the percentage of the general population that reports symptoms; (c) the relative likelihood of having Covid-19n for those with symptoms compared to those without symptoms; and (d) how more likely are people with symptoms to self-select into testing than those without symptoms. According to our methodology, it is possible to calculate these figures and thus estimate the bias. (a) is provided by the results of community testing; (b) is provided by surveying; (c) can be obtained by asking people a simple question before testing them for Covid-19; and (d) is provided by surveying.

A simple example is the following: Assume community testing led to 10% positive results, and 5% of the population reported symptoms. Without waiting time, if those with symptoms are 5 times more likely to have the virus than those without symptoms, then the results of community testing exaggerate by 27.71%, and the true prevalence in the population is 7.83% (instead of the reported 10%). At a 30-60 minute waiting time, the bias increases to 106.95%, meaning that the true prevalence in the population is 4.83%.

To further illustrate our results, Figure 1 depicts our best estimate of the virus prevalence bias, i.e. the ratio between reported prevalence and actual, depending on symptoms prevalence and waiting time, for the three age groups.

**Figure 1.**
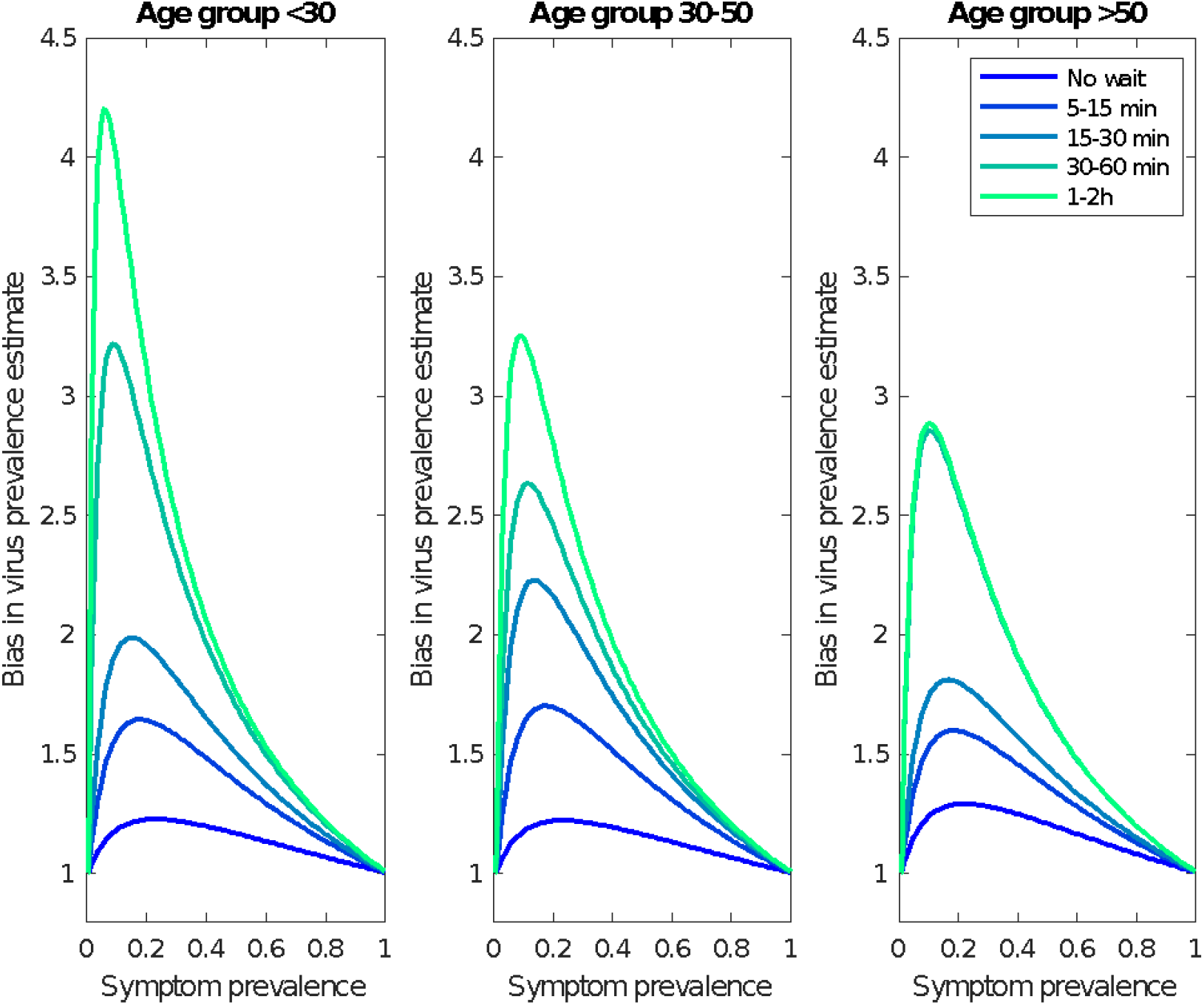
Best estimate of the virus prevalence bias: The ratio between reported prevalence and actual, depending on symptoms prevalence and waiting time, for the three age groups.

Based on these estimates, we can simulate how different demographic structures would affect the prevalence bias. In Figure 2 we depict the results from 3 million draws from the plausible parameter space (we assume symptoms prevalence of 5%, and allow the testing bias parameter to vary uniformly within the 95% confidence interval gained from the experiments) applied to three countries, with different demographic structures: Nigeria (with one of the youngest populations globally), Italy (heavily ageing population) and the USA (between the two extremes). The simulation shows that demography matters: a young country like Nigeria could have a substantially higher prevalence bias than Italy. However, it is also clear that the waiting times are more important than demographics. Lowering waiting times would result in a low bias for all countries.

**Figure 2.**
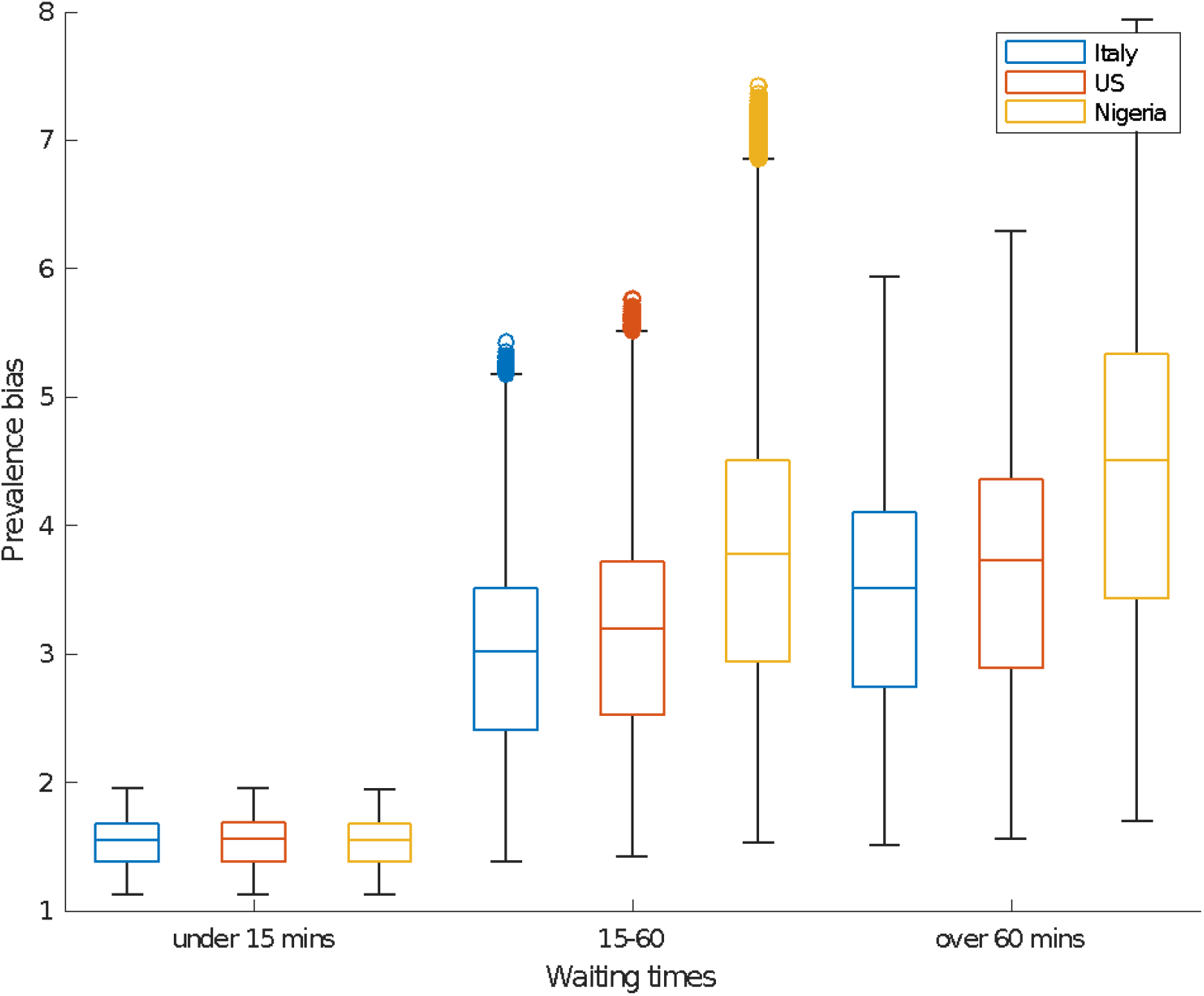
Simulation: How different demographic structures would affect the prevalence bias.

Finally, to validate our results on the prevalence bias, we compare with data from the REACT study in the UK. While in this study too, participants are freely choosing to test or not, testing kits are being sent home to participants at no cost and the sample is carefully selected. According to our results, the bias in this study is non negligible but should still be the smallest of any major field testing procedure yet published. Treating the (weighted) prevalence estimates from this study as true prevalence and comparing to the test positive rate from all tests done in the country in the same dates, we obtain an (independent to our experiments) estimate of the prevalence bias (see Appendix B). In the 10 different subwaves of the study, the estimated prevalence bias indeed is positive, substantial, but also highly variable, ranging from 3.8 to 23.6, thus confirming our three main predictions.

## 4. Discussion

Using an incentivised online experiment, we found that the probability of taking a Covid-19 test for those who have symptoms (or believe they are more likely to have caught the virus) is many times higher than those who do not. In our sample, this testing propensity bias ranged from 1.5 times (for people under 30 years with no waiting time) to 38 times (for people under 30 and a 2-hour waiting time). The bias becomes larger with longer waiting times, and any cost associated with taking the test. Testing stations cannot readily correct this by oversampling (i.e. selecting people without symptoms to test).

Demographics also influence the testing propensity bias, which means that different areas (or countries) will have different biases depending on the age composition. Furthermore, there have been reports of very long waiting times in community testing, which greatly exacerbates the bias. It is important to note that the bias is time-variant, and depends strongly on the actual virus prevalence.

Our findings imply that virus positivity results from community testing sites are heavily biased. Contrary to conventional wisdom in the health policy community that suggested the bias would be, if anything, downward, our results suggest that prevalence is inflated by up to 5 times, even if tests are not costly. The prevalence bias is not just large, but also time-varying, confirmed in a comparison with real-world data, meaning that test positivity comparisons across countries or time are necessarily flawed.

Importantly, the prevalence bias goes beyond the issues of age group or location selection usually considered. Rather, it relates to self-selection into testing for those who (believe they) are more likely to have Covid-19. This makes the aggregate results of community testing unreliable when it comes to drawing conclusions on the prevalence of Covid-19 in the population.

We recognise the importance of giving people the opportunity to test, as this identifies positive cases, thus allowing them to self-isolate and stop spreading the disease. If the goal of street testing is just to allow random people to have a quick and free test, then this possibly meets its goal. Note, however, that random testing is not efficient, economically, or epidemiologically: subsidising tests specifically for populations with a high risk of getting infected and infecting others would probably save more lives at lower cost (say, tests for young people working in service industries and living with their parents). These questions remain open for future research.

What we have shown is that “random” voluntary testing is not really random. As such, it does not provide accurate information on disease prevalence, which is important to design and implement urgent policy responses to the pandemic, in terms of type, intensity and geographic area. Since voluntary testing is always biased, aggregate results on prevalence *should* be corrected. Debiasing can be performed using our methodology, as long as there are good estimates for four parameters, namely (a) the percentage of tests in the field yielding positive results; (b) the percentage of the general population that reports symptoms; (c) how more likely are people with symptoms to be carrying the virus than those without symptoms; and (d) how more likely are people with symptoms to self-select into testing than those without symptoms. Obtaining estimates for the above parameters is of varying difficulty: (a) is obtained in any country doing “random” street testing, (b) can be estimated with standard polling and (d) can be estimated with our experimental methodology. Estimating (c) would require asking subjects at testing stations to self-report their symptoms before testing.

We suggest a novel, more economical and accurate alternative for prevalence estimation. The important parameter to estimate is the probability of having covid-19 conditional on having symptoms, and on not having symptoms, similar to parameter (c) above. This can be done by asking a simple question at existing testing sites (indeed we have ongoing parallel work underway to obtain these estimates in cooperation with testing centres). These parameters could be country-specific and time-variant, but we do not expect changes to be fast. Obtaining a few estimates in each virus season could suffice, and this estimate could be used for similar countries. The next step is unusual and often misunderstood: poll a representative sample regularly, *to obtain symptoms prevalence*. A common misunderstanding involves the argument that laymen cannot measure their symptoms properly. This is not a bug, but a feature of our procedure. Since the testing bias depends on self-reported symptoms, we need to condition on subjects *believing* they have symptoms, not on actually having them. Using both steps above can yield accurate prevalence estimates in real time at very low, comparatively, cost.

Our methodology is not limited to correcting the results of community testing. Confirmed cases reported daily are also biased, as some people might not test because of costs, or the inconvenience of going to a testing site, or even due to being afraid of losing income. According to our results, even at no monetary cost and no wait, 3.98% of people with symptoms would not get tested – which increases to 9.51% even for the slightest waiting time, rising even further when tests have a non-negligible cost to the citizen. Using polling results from a representative sample can correct this error.

The REACT study in the UK is an interesting special case of large-scale community testing on a nationally representative sample. The authors claim that this sample is truly random, but our results suggest this might not be the case. Even for people taking a free test at home (compare to the no waiting time condition in the experiment), a substantial testing bias exists. More importantly, REACT is very expensive to run, while simultaneously less timely than our polling proposal. While REACT has been done monthly or less often, our procedure can be run daily,

This paper also contributes to the literature on testing regimens.^15^ Mass testing, extending to a very large part of the population, is useful as it can provide more accurate figures, and also identifies positive cases. It has been used, among others, in Liverpool, Slovakia and South Korea.^16-19^ However, mass testing is extremely expensive, and might be infeasible, especially at frequent intervals, due to capacity and technical constraints.

In the absence of mass testing, obtaining unbiased prevalence estimates is of paramount importance for health and the economy. Underestimating disease prevalence can trigger inadequate measures and further spread of disease, while overestimating can be detrimental to economic activity. We thus urge policy makers to redesign “random” testing as a matter of priority in the effort to tackle the pandemic.

As a final note, our methodology is applicable to the prevalence measurement of *any* epidemic, when carriers have informative private information about their health status. Fighting disease is hard, even without the added complication of not knowing the location and magnitude of the fight. Our work offers tools to measure prevalence in real time. Further work is needed, to estimate specific selection-bias parameters for every disease, as they are necessarily related to the health burden and life expectancy reduction caused by the specific pathogen.

## Data Availability

The data used in this study were collected via surveys. We can make them available upon reasonable request

## Ethics approval

This study received ethics approval from the Economics Research Ethics Committee at City, University of London. Ethics approval was given on 9 December 2020. Code: ETH2021-0749.

## Data and materials availability

The data used in this study were collected via surveys.

## APPENDIX A

**Figure A1.**
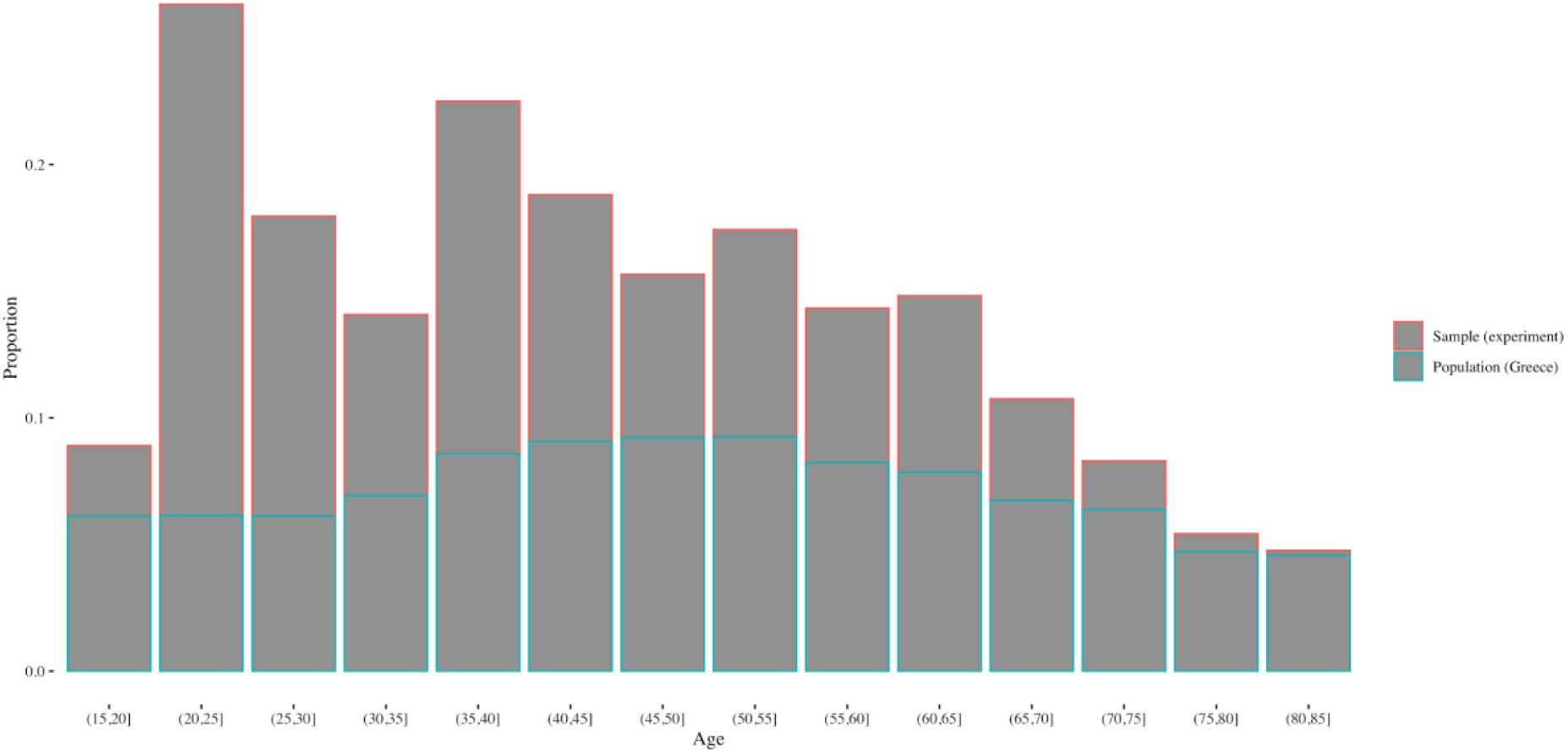
Age distribution in the experiment (n=578) and in population of Greece (source: populationpyramid.net).

**Figure A2.**
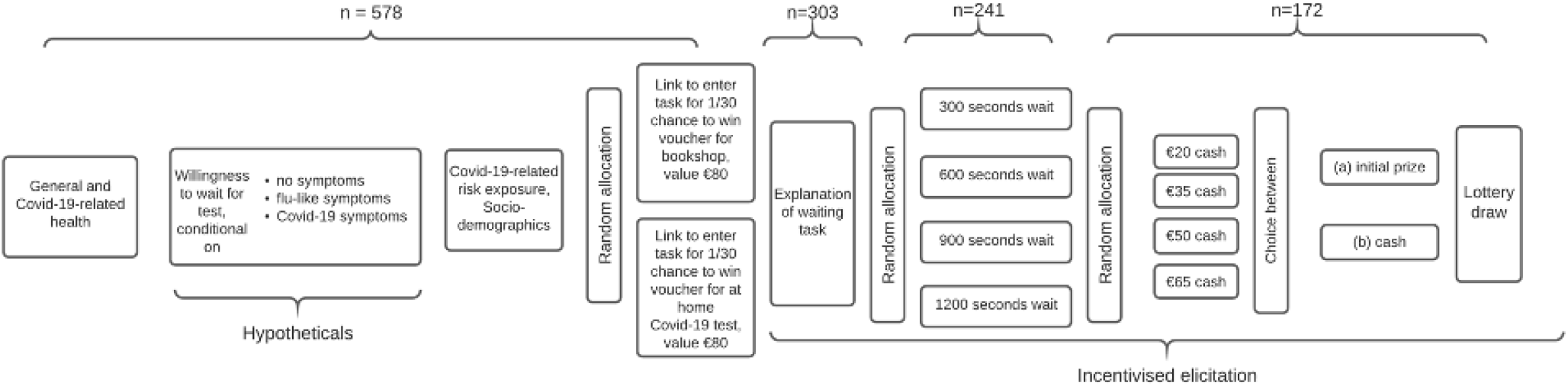
Flow of the experiment.

**Table A1.**
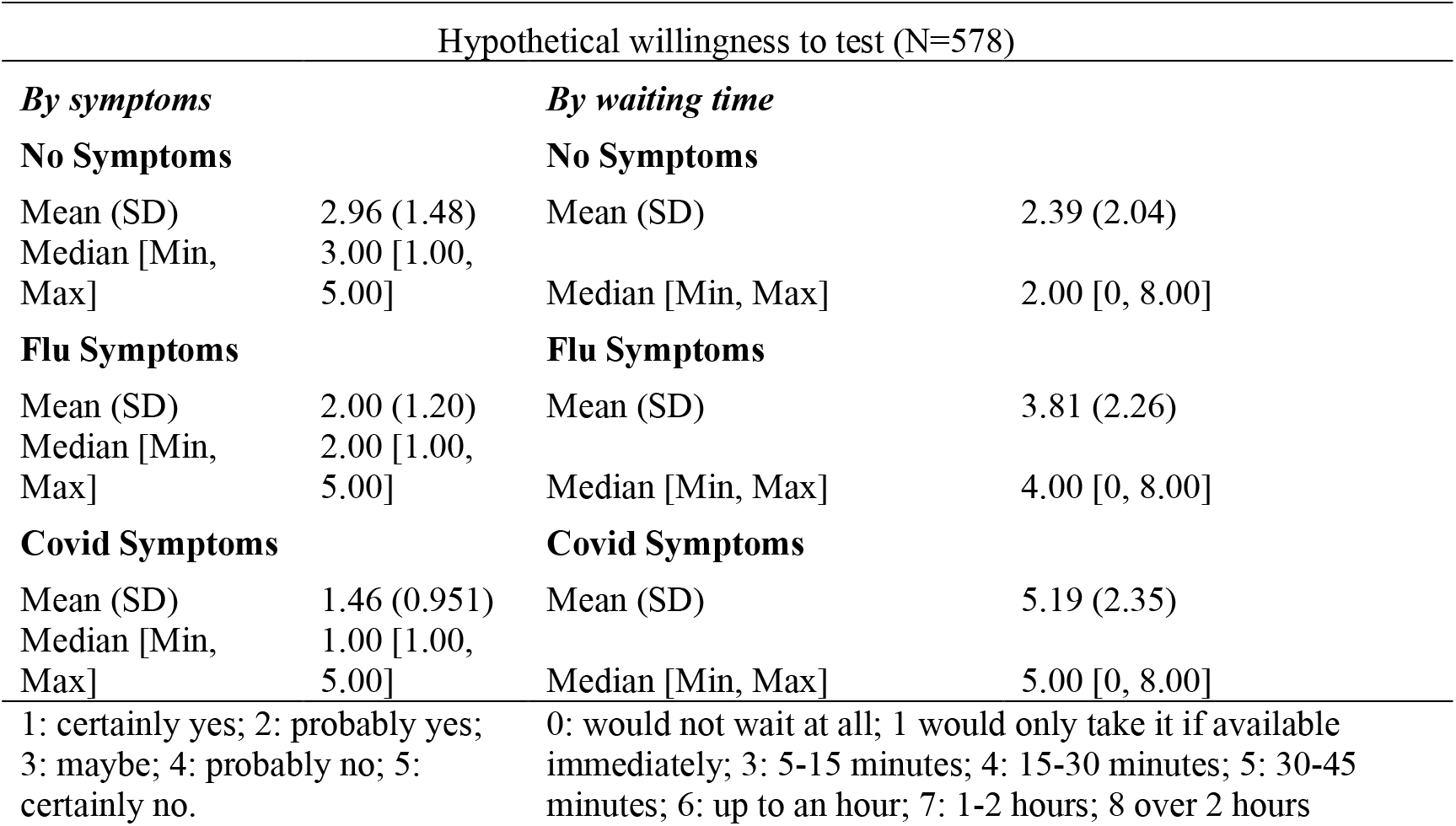
Summary Statistics

| Hypothetical willingness to test (N=578) |  |  |  |
| --- | --- | --- | --- |
| *By symptoms* |  | *By waiting time* |  |
| **No Symptoms** |  | **No Symptoms** |  |
| Mean (SD) | 2.96 (1.48) | Mean (SD) | 2.39 (2.04) |
| Median [Min, Max] | 3.00 [1.00, 5.00] | Median [Min, Max] | 2.00 [0, 8.00] |
| **Flu Symptoms** |  | **Flu Symptoms** |  |
| Mean (SD) | 2.00 (1.20) | Mean (SD) | 3.81 (2.26) |
| Median [Min, Max] | 2.00 [1.00, 5.00] | Median [Min, Max] | 4.00 [0, 8.00] |
| **Covid Symptoms** |  | **Covid Symptoms** |  |
| Mean (SD) | 1.46 (0.951) | Mean (SD) | 5.19 (2.35) |
| Median [Min, Max] | 1.00 [1.00, 5.00] | Median [Min, Max] | 5.00 [0, 8.00] |
| 1: certainly yes; 2: probably yes; 3: maybe; 4: probably no; 5: certainly no. |  | 0: would not wait at all; 1 would only take it if available immediately; 3: 5-15 minutes; 4: 15-30 minutes; 5: 30-45 minutes; 6: up to an hour; 7: 1-2 hours; 8 over 2 hours |  |

**Table A2.** Willingness to wait for a 1/30 chance of winning a prize. Number of subjects by level of task completion and incentive (rows 1-2), bias by incentive (row 3).

| Prize | Not entered | Dropped upon learning waiting time | Dropped after some wait | Swapped prize for cash | Kept prize | N |
| --- | --- | --- | --- | --- | --- | --- |
| **Book voucher** | 103 | 31 | 38 | 46 | 48 | 266 |
| **Test voucher** | 138 | 25 | 31 | 62 | 12 | 268 |
| **Bias** | 1,263 | 1,267 | 1.28 | 4.03 | 0.25 | 534 |

## APPENDIX B

The REACT study has been conducted in eight waves, to date. Two of the waves have been published in two subwaves, yielding 10 different observations. We focus on the weighted prevalence figure published, as the most accurate. The number of daily tests is publicly available, along with the number of tests being positive, yielding test positivity. We divide test positivity by the REACT prevalence estimate to obtain an estimate of the prevalence bias in field testing. The following graph presents the results.

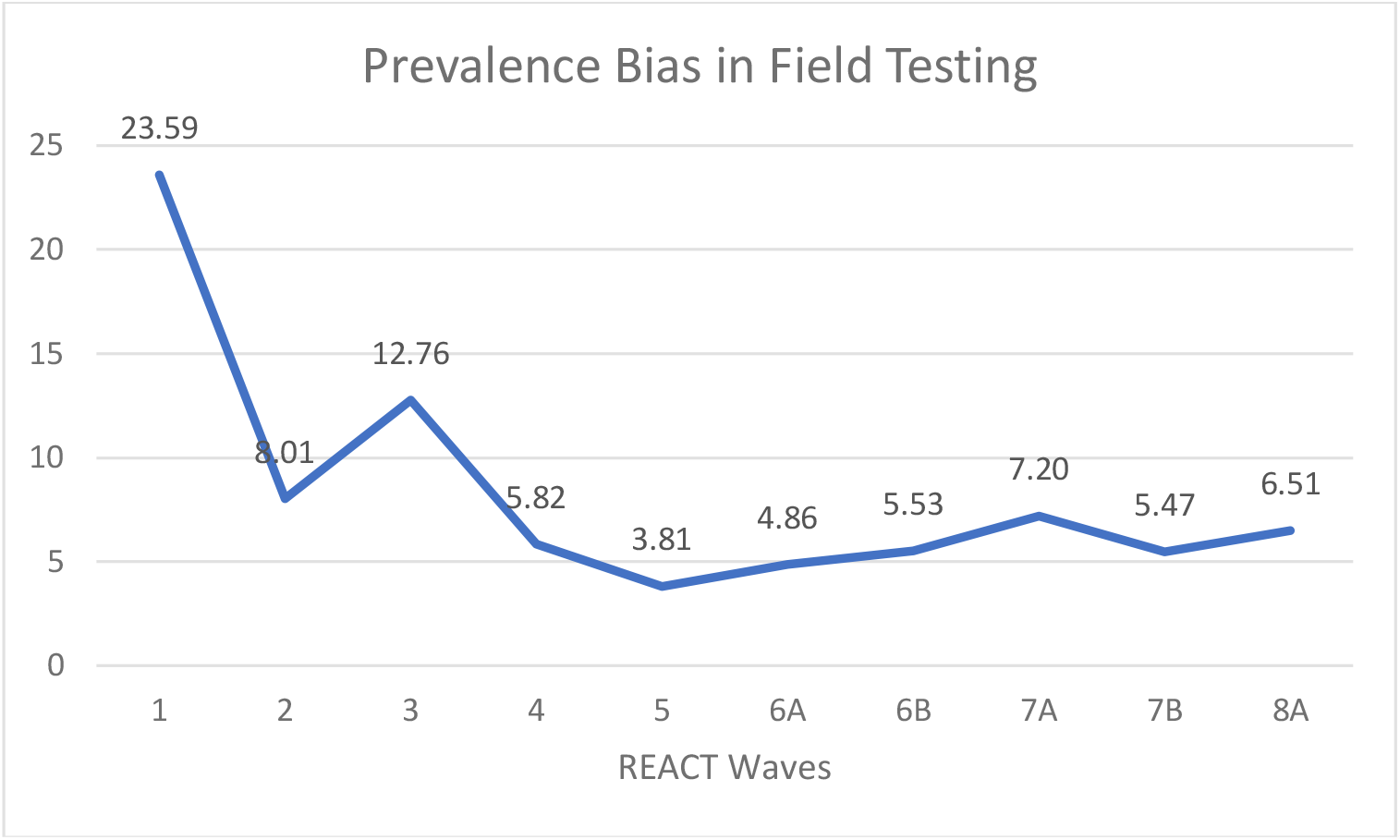

We then compare prevalence estimates using several different methods and three sources of data. The REACT data consists of non-overlapping random samples of the population of England at lower-tier local authority level that were invited to take part in each round of the study based on the National Health Service list of patients. The Office for National Statistics (ONS) data from Covid-19 Infection Survey (available at: https://www.ons.gov.uk/peoplepopulationandcommunity/healthandsocialcare/conditionsanddiseases/datasets/coronaviruscovid19infectionsurveydata) is a combination of cross-sectional and (where consent was obtained) repeated visits to the UK households. For comparability, we restricted the sample to data on England only. The Our World in Data (https://ourworldindata.org/) estimates reflect the total test positivity for the country’s sources.

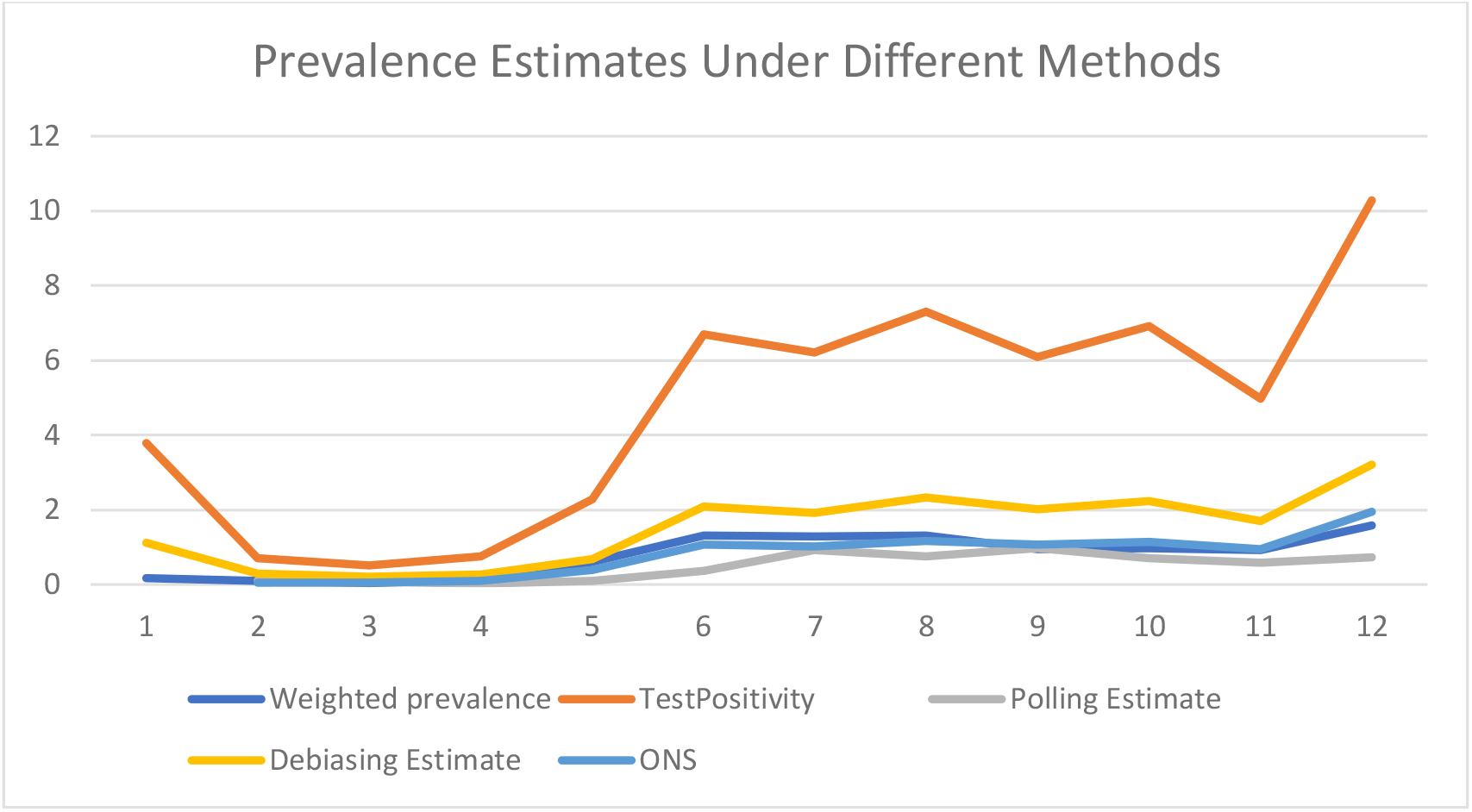

The above graph shows prevalence estimates (in percentage points) over the 12 month of 2020. The blue line depict the ONS prevalence estimate, the orange line tracks the OWD estimate, and the remaining lines reflect different methods of prevalence estimation applied to the REACT data. First thing to note is that the REACT weighted prevalence estimate, the ONS prevalence estimate and the polling estimate we compute using the current symptoms estimates but with the previous waves virus positivity by symptoms – track each other fairly closely. Secondly, these results are strikingly at odds with the overall OWD estimate. This is in line with our predictions of different testing approaches leading to different size of the bias.

## APPENDIX C

### Bias calculations

The aim is to infer the percentage of sick people in the population from the “random” testing in the field figures, as released by several Health Agencies worldwide. The problem is that testing is voluntary, which leads to selection bias. How large is this bias?

To start, some people believe they have symptoms, some don’t: call them S(ymptomatic) and H(ealthy). Note that the discussion below has to do with what people believe, not what they actually have. Also, we distinguish between people believing they have symptoms and those who do not, but the analysis readily extends to people having strong beliefs that they might be carrying the virus and those who do not.

Let the frequency of people who believe they have symptoms be p_s_, or just p, with 1-p being the frequency of people who do not think they have symptoms.

Of each group, some percentage turns out having the virus. Let v_s_ be the virus prevalence for those who believe they have symptoms, v_h_ for those who do not.

Of each group, some percentage are willing to take the test (for a given waiting time to take the test). Assume this only depends on symptoms, but not on actually having the virus (this assumption is mostly innocuous, unless there is a very large number of people in hospital). Let then t_s_ be the percentage of people who believe they have symptoms who actually take the test, and t_h_ for those who do not.

True prevalence is then

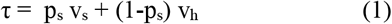

Given parameters, what number shows up positive in the sample (assuming that the test itself is perfect)

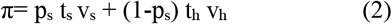

Divide by the total sampling rate

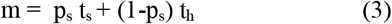

to get the sample prevalence (or virus frequency in the sample population) φ

Note that if t_s_=t_h_=t, then π = t (p_s_ v_s_ + (1-p_s_) v_h_) and φ = t (p_s_ v_s_ + (1-p_s_) v_h_)/t = p_s_ v_s_ + (1-p_s_) v_h_ =τ which makes sense; if testing propensities are equal, there is no bias.

If on the other hand the testing propensities t are not the same, then the sample is selected, leading to bias. Before we calculate the bias, express the propensities to test and be virus positive, for the people who believe they have symptoms, as a multiple of the propensities of those who do not: v_s_ = a v_h_, t_s_ = b t_h_.

Using these equations, rewrite (1), (2) and (3).

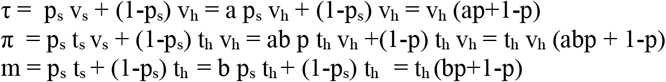

Simplify notation by writing p for p_s_ and calculate

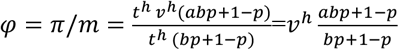

Now, to find the size of the bias, divide φ/τ

v_h_ (abp + 1-p) / (bp+1-p) / v_h_ (ap+1-p)

⇨ The bias in prevalence estimates β is

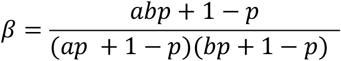

### Examples

Suppose a=1

β=(bp+1-p) /(bp1-p))/(a p + 1 – p)= 1/(p+1–p)=1

So, both a and b are necessary for the bias to exist, which makes sense conceptually.

Suppose a=b>1

Conceptually it is not unlikely that the two propensities be of similar magnitude, since the higher the risk of carrying the virus conditional on having symptoms, the more likely it should be people with symptoms seek testing.

The bias becomes

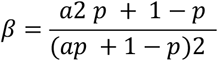

Let a=b=10

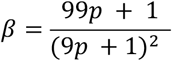

This means β=3 at p=0.1, at p=0.05 we still get β=2.7

Assume unequal a and b, but p=0.1. Then

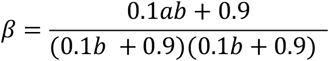

This function is plotted in the next graph, with a in the x axis, and b in the y axis.

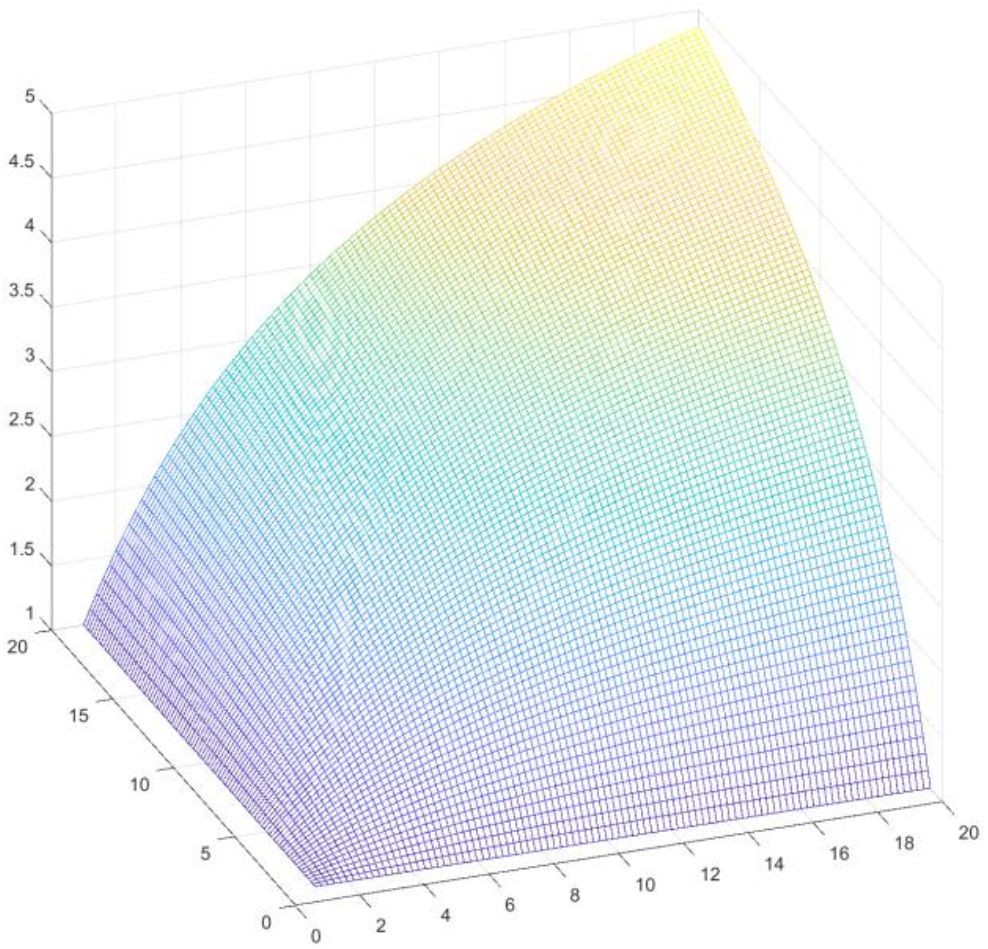

For a=b=20, street testing is overestimating the virus prevalence about 5 times.

#### Getting p_s_ from φ

While we suggest to get p_s_ through random (unbiased) polling of people about their perceived symptoms, it is also possible to calculate it using a, b, v_h_ and φ as follows.

Start with the definition of φ=v_h_ (abp + 1-p) / (bp+1-p)

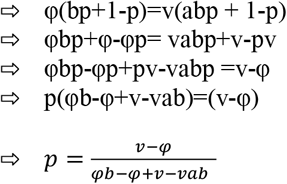

Note:

The denominator is negative φb-φ+v-vab<0

=> φ(b-1)>v(1-ab), since v(1-ab) is negative

=> φ>v(1-ab)/(b-1)

Which is true since b-1 is positive.

For v<φ the numerator is also negative, meaning p is positive.

If φ= v_h_ then symptoms prevalence is 0, all people in the sample have no symptoms, and v_h_ show positive in the test.

If φ= v_s_ =av_h_ then (v-av)/(avb-av+v-vab)=1, meaning everyone has symptoms, p=1.

Obviously φ cannot be above v_s_ (sample prevalence is highest if you only have people with symptoms in the sample, in which case not more than v_s_ can be positive)!

So we can debias the health agencies’ numbers without knowing p_s_

Again, it is easier not to do street testing, but to use v_h_ and v_s_ and poll about p.

### Examples

Suppose a=10, b=3 and v_h_=0.01

⇨ p=(0.01-φ)/(3φ-φ+0.01-0.3) = (0.01-φ)/(2φ-0.29)

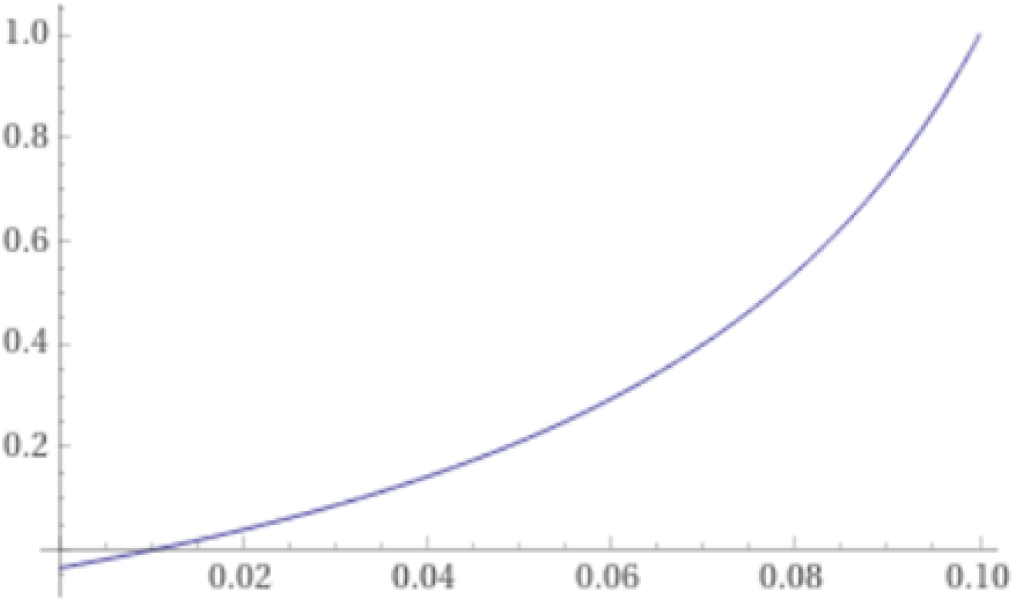

So, for example φ=0.1 yields p=-0.09/-0.09=1.

This makes sense. Everyone had symptoms, and v_s_=0.1 means that 10% had the virus, which is the proportion you will find in any sample. The interpretation is that with such a low true virus prevalence, the only way to get a relatively high φ is if there are *only* symptomatic people.

Suppose a=10, b=3 and v_h_=0.1

P=(0.1-φ)/(3φ-φ+0.1-3)= (0.1-φ)/(2φ-2.9)

So, in this case, p is about half φ for many values of φ.

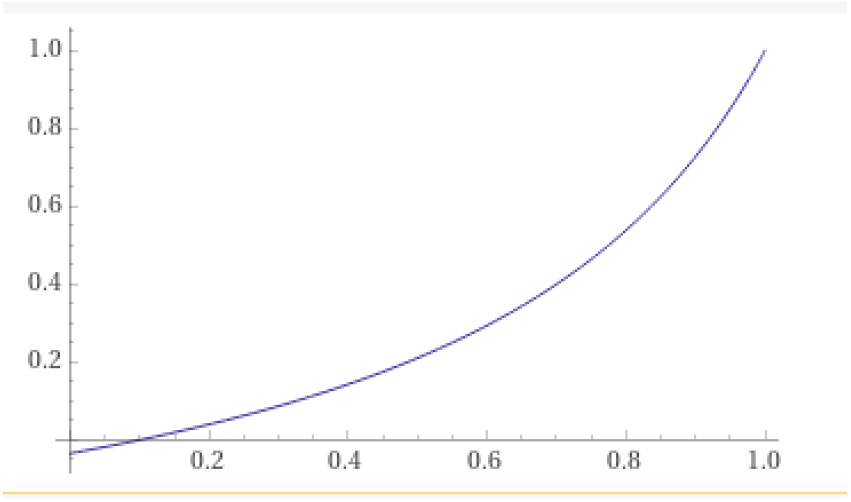

Now, let a=3, b=10 and v_h_=0.1

The effect of a and b is not symmetric.

p=(0.1-φ)/(10φ-φ-2.9)

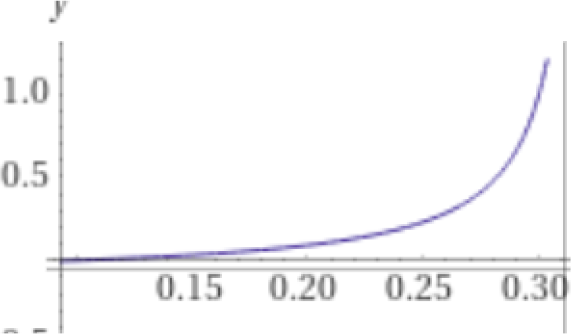

#### Getting p_s_ from biased symptoms prevalence in the test

Suppose that some national agency is asking about (perceived) symptoms before testing. It is then easier to debias and find the symptom prevalence in the general population p_s_

Symptom prevalence in the test would be

χ= pt_s_ / (pt_s_ + (1-p)t_h_)= bpt_h_ / (bpt_h_ + (1-p)t_h_)

_=>_ χ=bp/(bp + 1-p)

So true symptom prevalence is

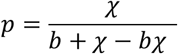

For a relatively low b=3

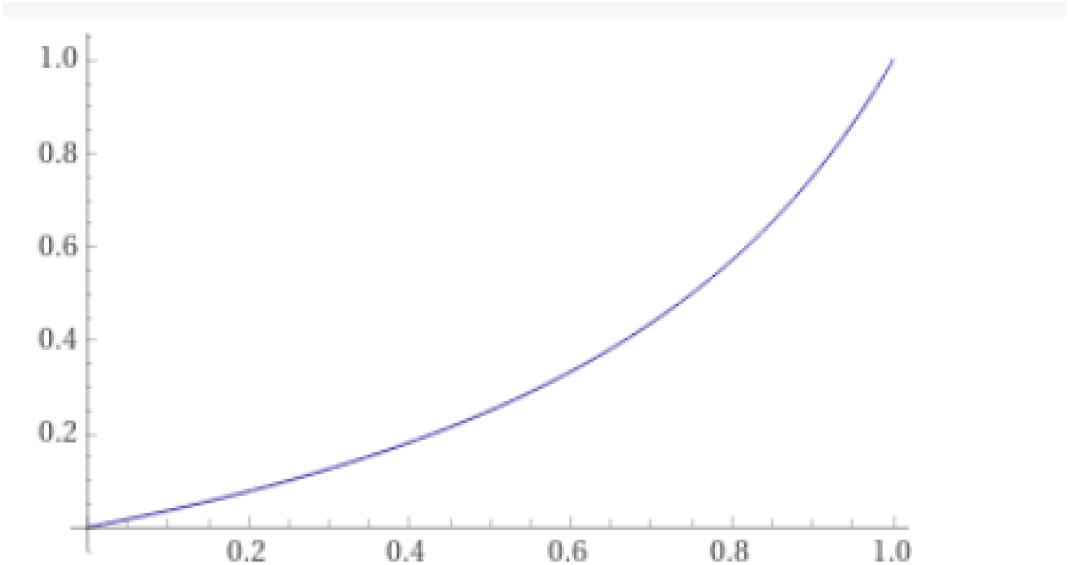

## References

[1] Vandoros, S., 2020. Excess Mortality during the Covid-19 pandemic: Early evidence from England and Wales. Social Science & Medicine, p.113101.

[2] Maringe, C., Spicer, J., Morris, M., Purushotham, A., Nolte, E., Sullivan, R., Rachet, B. and Aggarwal, A., 2020. The impact of the COVID-19 pandemic on cancer deaths due to delays in diagnosis in England, UK: a national, population-based, modelling study. The Lancet Oncology, 21(8), pp.1023–1034.

[3] Solomon, M.D., McNulty, E.J., Rana, J.S., Leong, T.K., Lee, C., Sung, S.H., Ambrosy, A.P., Sidney, S. and Go, A.S., 2020. The Covid-19 Pandemic and the Incidence of Acute Myocardial Infarction. New England Journal of Medicine.

[4] Ueda, M., Nordström, R. and Matsubayashi, T., 2020. Suicide and mental health during the COVID-19 pandemic in Japan. medRxiv.

[5] Tanaka, T. and Okamoto, S., 2020. Suicide during the COVID-19 pandemic in Japan. medRxiv.

[6] Chudik A, Mohaddes K, Perasan MH, Raissi M, Rebucci A. 2020. Economic consequences of Covid-19: A counterfactual multi-country analysis. VoxEU. Available at: https://voxeu.org/article/economic-consequences-covid-19-multi-country-analysis

[7] McInerney, M. and Mellor, J.M., 2012. Recessions and seniors’ health, health behaviors, and healthcare use: Analysis of the Medicare Current Beneficiary Survey. Journal of Health Economics, 31(5), pp.744–751.

[8] Riumallo-Herl, C., Basu, S., Stuckler, D., Courtin, E. and Avendano, M., 2014. Job loss, wealth and depression during the Great Recession in the USA and Europe. International Journal of Epidemiology, 43(5), pp.1508–1517.

[9] Thornton, R., 2008. The Demand for, and Impact of, Learning HIV Status. American Economic Review. 98(5). 1829–63

[10] Riley et al. 2021. REACT-1 round 8 interim report: SARS-CoV-2 prevalence during the initial stages of the third national lockdown in England, working paper.

[11] Atkeson, A., 2020. How deadly is covid-19? understanding the difficulties with estimation of its fatality rate (No. w26965). National Bureau of Economic Research.

[12] ECDC, 2020. COVID-19 testing strategies and objectives Available at: https://www.ecdc.europa.eu/sites/default/files/documents/TestingStrategy_Objective-Sept-2020.pdf

[13] ECDC, 2021. Data on testing for Covid-19 by week and country. Available at: https://www.ecdc.europa.eu/en/publications-data/covid-19-testing

[14] Georganas S., Laliotis, I., and Velias A. 2021. The Best is Yet to Come: The Impact of Retirement on Prosocial Behavior, working paper

[15] Mina, M.J., Parker, R. and Larremore, D., 2020. Rethinking Covid-19 Test Sensitivity - A Strategy for Containment New England Journal of Medicine, 383:e120

[16] Pavelka, M., van Zandvoort, K., Abbott, S., Sherratt, K., Majdan, M., Jarcuska, P., Krajci, M., Flasche, S., Funk, S. and CMMID COVID-19 working group, 2020. The effectiveness of population-wide, rapid antigen test based screening in reducing SARS-CoV-2 infection prevalence in Slovakia. medRxiv.

[17] BBC, 2020. Covid: Mass testing in Liverpool sees ‘remarkable decline’ in cases. Available at: https://www.bbc.com/news/uk-england-merseyside-55044488 Accessed 19 December 2020.

[18] Bloomberg, 2020. Seoul’s full cafes, apple store lines and show mass testing success. Available at: https://www.bloomberg.com/news/articles/2020-04-18/seoul-s-full-cafes-apple-store-lines-show-mass-testing-success Accessed 19 December 2020.

[19] Brauner et al., 2020, Inferring the effectiveness of government interventions against COVID-19. Science, forthcoming

